# Testing lags and emerging COVID-19 outbreaks in federal penitentiaries: A view from Canada

**DOI:** 10.1101/2020.05.02.20086314

**Authors:** Alexandra Blair, Abtin Parnia, Arjumand Siddiqi

## Abstract

**Objectives:** To provide the first known comprehensive analysis of COVID-19 outcomes in a federal penitentiary system. We examined the following COVID-19 outcomes within federal penitentiaries in Canada and contrasted them with estimates for the overall population in the penitentiaries’ respective provincial jurisdictions: testing, prevalence, the proportion recovered, and fatality.

**Methods:** Data for prisons were obtained from the Correctional Service of Canada and, for the general population, from the Esri COVID-19 Canadian Outbreak Tracking Hub. Data were retrieved between March 30 and April 21, 2020, and are accurate to this date. Penitentiary-, province- and sex-specific frequency statistics for each outcome were calculated.

**Results:** Data on 50 of 51 penitentiaries (98%) were available. Of these, 72% of penitentiaries reported fewer tests per 1000 population than the Canadian general population average (16 tests/1000 population), and 24% of penitentiaries reported zero tests. Penitentiaries with high levels of testing were those that already had elevated COVID-19 prevalence. Five penitentiaries reported an outbreak (at least one case). Hardest hit penitentiaries were those in Quebec, Ontario, and British Columbia, with some prisons reporting COVID-19 prevalence of 30% to 40%. Of these, two were women’s prisons. Female prisoners were over-represented among cases (31% of cases overall, despite representing 5% of the total prison population).

**Conclusion:** Increased sentinel or universal testing may be appropriate given the confined nature of prison populations. This, along with rigorous infection prevention control practices and the potential release of prisoners, will be needed to curb current outbreaks and those likely to come.

**GRAPHICAL SUMMARY:** 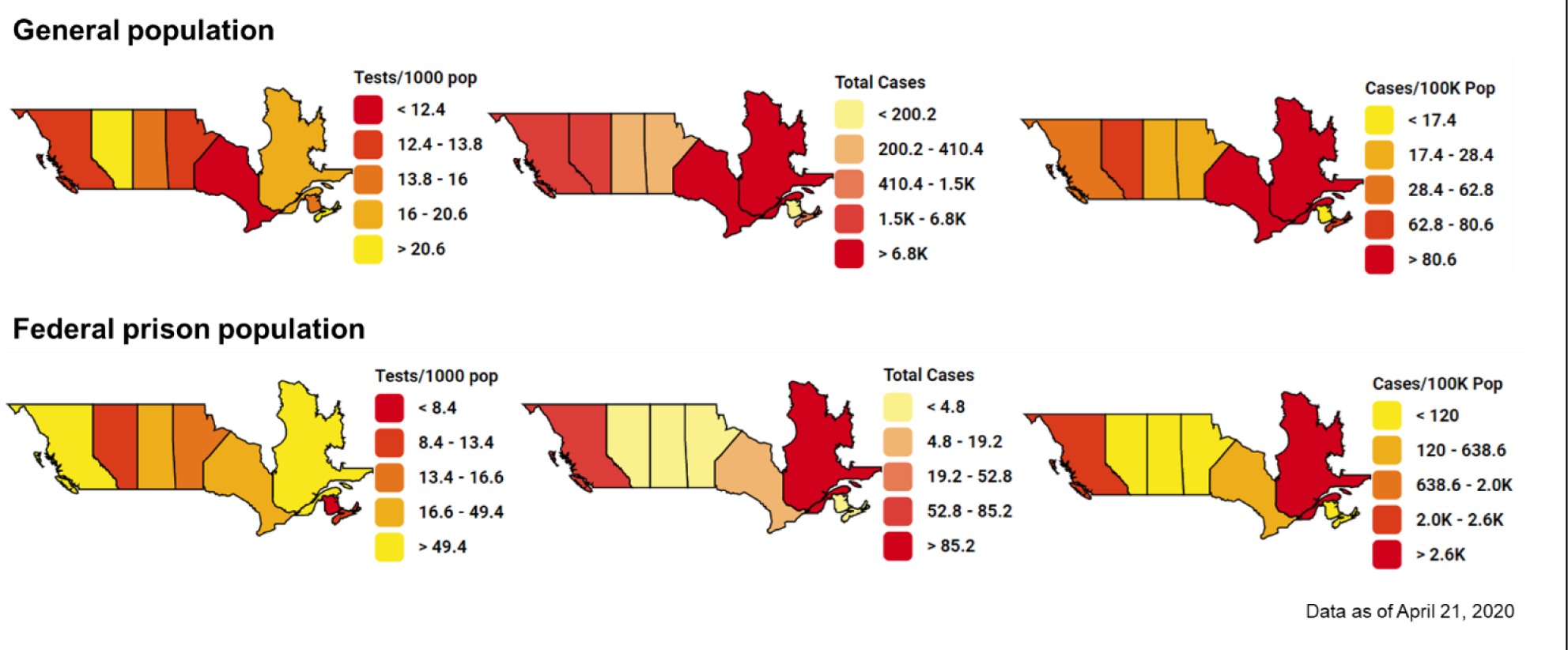

- Between 20% and 57% fewer tests per 1000 population have been conducted in federal prisons in Saskatchewan, New Brunswick, Nova Scotia and Alberta than in the general population of those provinces.
- Though Alberta, Manitoba, Saskatchewan, New Brunswick and Nova Scotia are reporting lower counts of COVID-19 cases, these are also the regions reporting the lowest levels of testing.
- Case incidence has been highest in federal prisons in Quebec, Ontario, and British Columbia, where a total of five prisons are experiencing outbreaks (1 or more cases). These regions are those reporting the highest levels of testing – higher than the testing levels in the general population.

## 1. BACKGROUND

As we are (re)learning from the evidence of COVID-19 outbreaks in long-term care facilities, housing large groups of individuals in confined, institutionalized environments is a recipe for large-scale, human-to-human transmission of infectious disease. Prisons have similar dynamics of confinement, of residents who are dependent on the institutions’ conditions and policies for basic living and hygiene and, of clinical susceptibility due to underlying health conditions [1–3]. Epidemiologic findings from past outbreaks of respiratory diseases such as influenza, adenoviruses, and tuberculosis in high-income countries highlight the higher likelihood of infection transmission and disease incidence within prisons compared to the general population. [1–3]. Early reports suggest that several penitentiaries in the United States are experiencing COVID-19 outbreaks [4], though, as far as we know, no comprehensive analysis of theses penitentiaries has yet been conducted, and very little is known about COVID-19 in prison contexts in other high-income countries. In this study, we use Canadian data to provide the first known examination of COVID-19 statistics for a federal penitentiary system. In doing so, we also bring front-and-center a major health equity issue, because Indigenous and racialized communities are over-represented within the Canadian prison system [5].

In the context of the COVID-19 pandemic, there are specific factors that place prisoner populations at particularly high risk of COVID-19 infection, and of a higher risk for severe (compared to mild) COVID-19 outcomes. These include housing multiple prisoners to one cell (“double-bunking”)[6], the ageing population within prisons—particularly in federal prisons[7], a high prevalence of chronic disease comorbidities and immuno-compromised health status associated with substance use and blood-borne infections[8], and the regular entrance of custodial and health care staff from communities with likely community-based transmission.

At the time of writing (late April 2020), Canada has been documenting the COVID-19 pandemic on its soil for almost two months. In late March 2020, the Correctional Service of Canada (CSC), Canada’s federal body responsible for incarcerated individuals serving sentences of at least two years, had indicated that they had taken steps to prevent and were prepared to respond to COVID-19 outbreaks inside its federal penitentiaries [9]. With this study, we aim to offer a baseline situational assessment of COVID-19 testing and burden in Canadian federal penitentiaries, from which future analyses can be based. This study aims to summarize and compare the prevalence of testing, test positivity, COVID-19 prevalence, case fatality, and the proportion of cases recovered within federal penitentiaries in Canada, by province and for Canada overall, and to contrast these with rates in penitentiaries’ respective provincial jurisdictions.

## 2. METHOD

### Data and study population

In an unprecedented act of transparency, CSC made COVID-19 testing and case numbers publicly available for each federal penitentiary on their website. Data up until April 21, 2020, were used [10]. Equivalent values for the Canadian population were extracted from the COVID-19 Canada Outbreak Tracker Resource Hub [11]. For each indicator, described below, we calculated prison-, sex-, and province-specific estimates. Prison population denominators were approximated by the maximum prison capacity for each penitentiary [12]. For the general population, denominators (i.e., total general population) were obtained from Statistics Canada population estimates for the first quarter of 2020 [13].

Additionally, to provide a timeline for the number of tests and cases in prisons with possible outbreaks (5 or more cases at the time of analysis), we used the Wayback Machine (archive.org) to obtain all previous copies of CSC’s COVID-19 reporting webpage [10]. The earliest version of the CSC webpage was created on April 1, 2020, which provided cumulative data up until March 30, 2020. There were eight additional updates, which reported on data from April 7, 8,10, 12, 13, 15, 17, and 18, 2020. Cumulative data at each of these dates were merged with the data from April 21, 2020, to create a time-series ranging from March 30 to April 21, 2020. For calendar dates at which no CSC updates were made available, we filled in the information with the data from the previous observation.

Given that several federal penitentiaries are in fact multiplex sites—either with distinct buildings or units operating under different security levels (e.g. minimum, medium or maximum) or offering distinct services (e.g. treatment facilities)—and that sub-population capacities were not always available for each separate unit, we chose to group several multi-complex facilities in our analyses. This was the case of Quebec’s Federal Training Center Multi-Level Unit and Minimum security facility; British Columbia’s Pacific Institution, Pacific Institution Regional Treatment Center and Pacific Institution Reception Center; Ontario’s Millhaven Institution, Millhaven Regional Hospital and Millhaven Regional Treatment Center, as well as Collins Bay Minimum Institution and Collins Bay Regional Treatment Center, and Joyceville Institution and Joyceville Minimum Institution. With these groupings, we recorded 51 facilities. We were able to analyze data from 50 of these facilities (98%) (all data in Supplementary File 1).

### Measures

#### Total tests and cases

The total number of tests performed was obtained directly from the CSC and the Canada Outbreak Tracker Resource Hub websites. CSC reports on the number of “positive tests”. We considered all “positive tests” as confirmed cases.

#### Tests per 1000 population

Tests per 1000 population were estimated by dividing the total number of tests performed by the total population in each facility, in the prisoner population of each province, and the general population of each province, respectively, and multiplying the fraction by 1000.

#### Test-positive rate

We estimated the proportion of tests that were positive by dividing the total number of positive tests (confirmed cases) by the total number of tests, in each prison, provincial prison population, and the provincial general population.

#### Tested COVID-19 prevalence

We estimated the prevalence of COVID-19 as identified through tests in the penitentiary and provincial populations by dividing the total number of positive tests (confirmed cases) by the population of each facility, the total prison population by province, and the total provincial population, respectively. This indicator can also be considered the cumulative incidence proportion.

#### Proportion recovered

To offer a crude estimate of the evolution of the epidemics inside and outside of prisons, we estimated the proportion of cases that had recovered. This was done by dividing the number of cases who had recovered by the total number of positive tests (total confirmed cases).

Operational definitions of recovered cases vary across jurisdictions in Canada, and CSC does not define their measure in their reporting. Recovered cases are those who either received confirmatory negative test results or for whom 10 to 14 days have elapsed since the start of their symptoms and they are symptom-free for at least 2 to 3 days [14, 15]. Therefore, we interpret recovered case numbers with caution.

#### Population categories in prisons with outbreaks: Susceptible, Infected, Recovered and Died

An outbreak of COVID-19 within confined populations, such as long-term care homes, can be declared after a single resident or staff member is confirmed positive [16]. Thus, we consider prisons with one or more reported cases of COVID-19 among its prisoner population as those experiencing outbreaks. To describe the evolution of testing and incidence in prisons with outbreaks across our study period, we classified each prison’s population into four categories, for each calendar day of the study period. We estimated the number of prisoners who were “susceptible” to infection by subtracting the total number of confirmed active, recovered, and deceased cases from the maximum population capacity of each prison. Prisoners considered “infected” were those with positive tests who had yet to recover or die. Prisoners declared to be “recovered” were assumed to no longer present active infections or be susceptible to infection. Lastly, the total number of prisoners who had “died” were those whose death was declared to result from COVID-19 complications.

## 3. RESULTS

### 3.1 Testing inside versus outside federal prisons

An analysis of testing within all federal penitentiaries illustrates both the heterogeneity of COVID-19 testing within prisons in Canada and a low if not a complete absence of testing in many facilities (Figure 1). Twelve of the 50 facilities studied—nearly one in five (24%)—had recorded a complete absence of testing (indicated by hollow bars in Figure 1). Approximately 72% of all facilities (36/50) recorded fewer tests than the Canadian general population average of 16 tests per 1000 population. Facilities with higher levels of testing per 1000 population, tended to be those that had already reported a higher COVID-19 prevalence (Figure 1).

**Figure 1:**
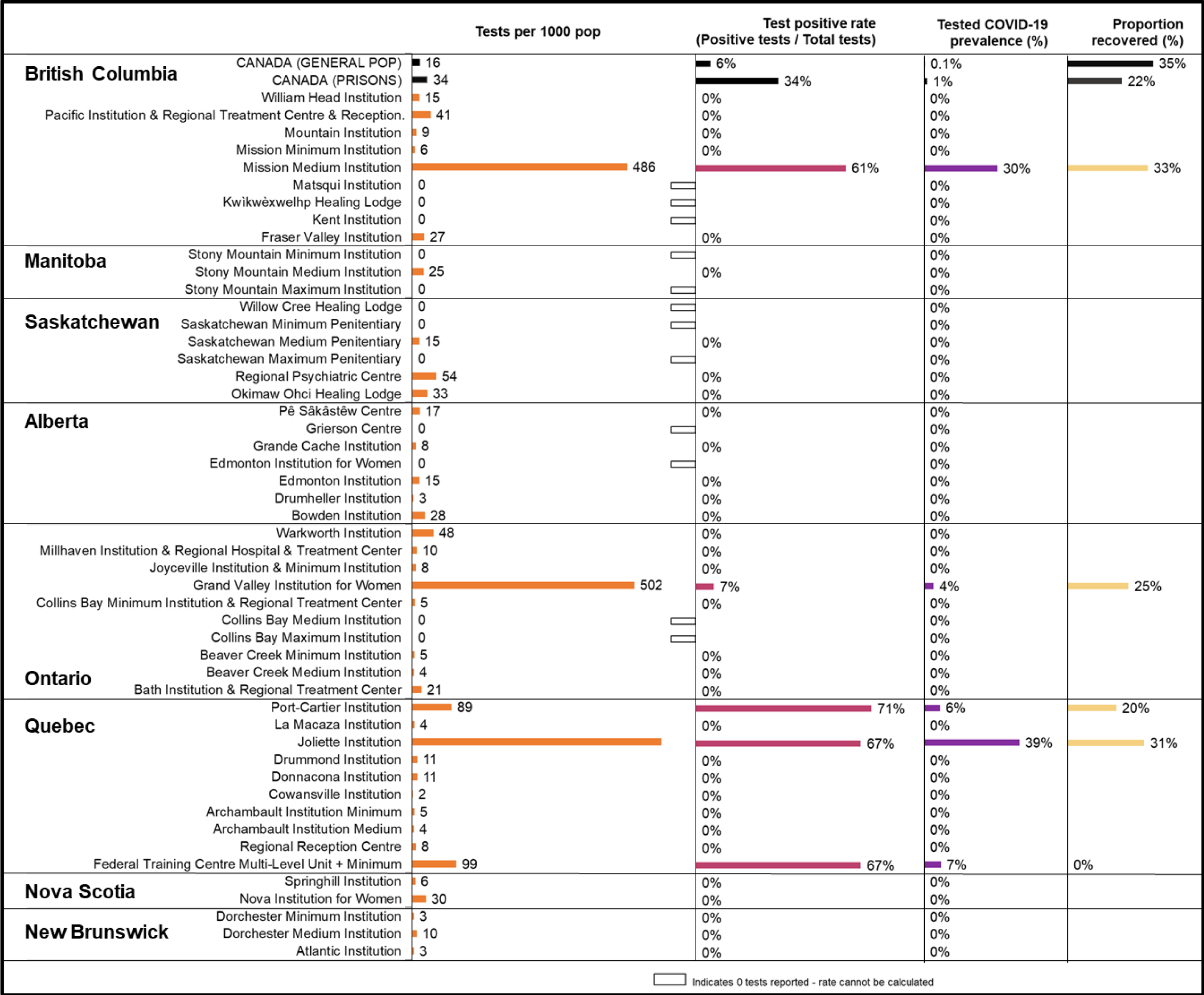
Test-positive proportion, case prevalence, and testing per 1000 population for all federal penitentiaries.

On average across provinces, New Brunswick, Nova Scotia, Alberta, and Saskatchewan have been testing less inside prisons than in the general population (Figure 2). The opposite is true for Quebec, Ontario, and British Columbia, which were likely responsible for driving the average of 34 tests per 1000 population observed for the prison population, overall.

**Figure 2:**
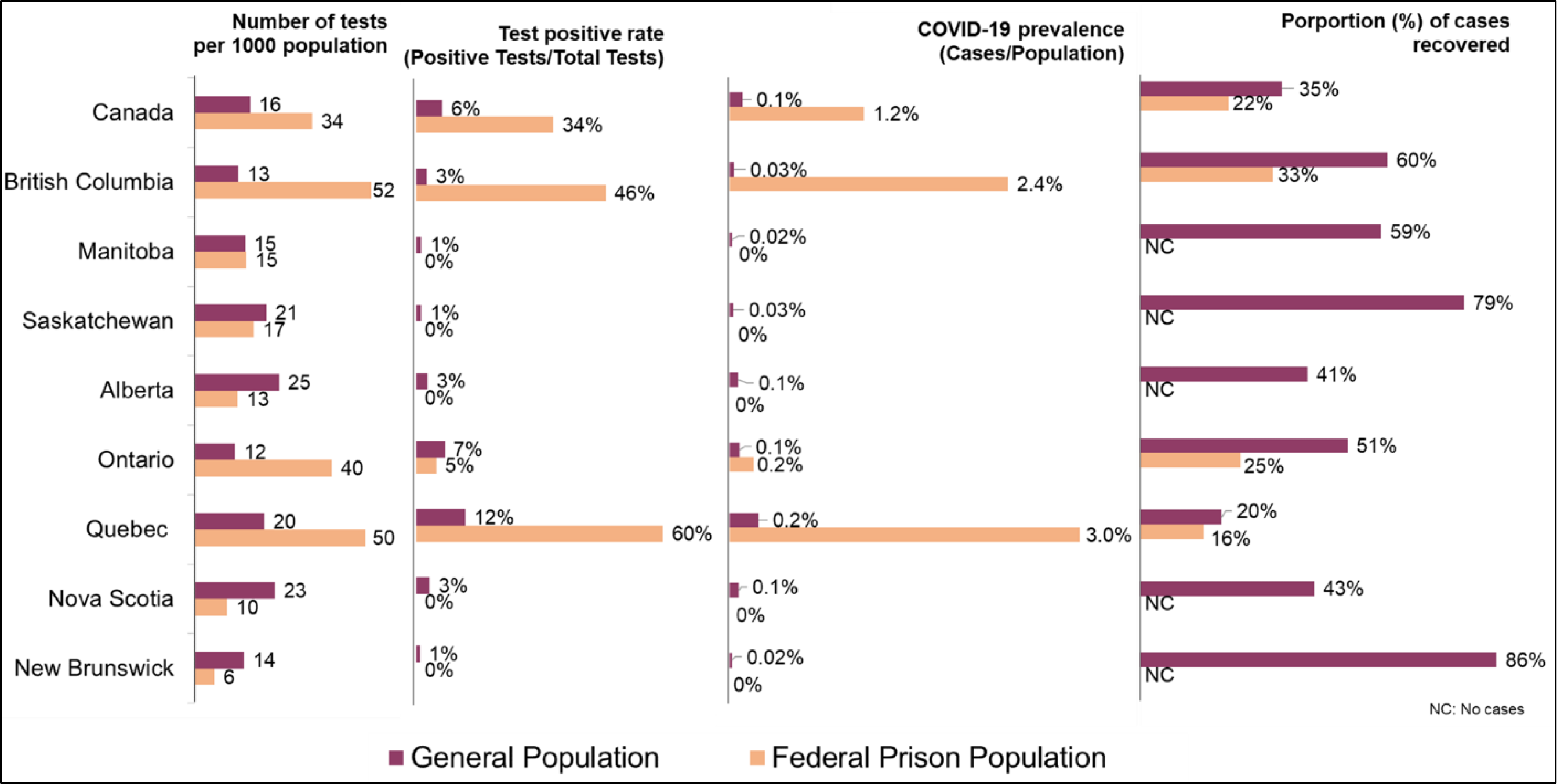
Testing per 1000 population, test-positive rate, and prevalence of positive tests, for federal prison and general populations, by province and in Canada overall

### 3.2 Prevalence of COVID-19 inside versus outside federal prisons

The high positive test rates and estimated prevalence of COVID-19 in prisons in Quebec and British Columbia point to emerging outbreaks in these provinces (Figure 2). Prevalence was 10 times higher in Quebec, 6 times higher in British Columbia, and 2 times higher in Ontario within federal prisons compared to the general population of each province (Figure 2).

Many of the hardest-hit penitentiaries are near major city centers. In Quebec, this was the Federal Training Center complex in Laval, just north of Montreal (51 cases, 7% COVID-19 prevalence) and, Joliette Institution for Women in Joliette, a 1-hour drive away from Montreal (51 cases, 39% prevalence). However, Port Cartier Institution in Côte Nord (a relatively remote region of the province) also reported 15 cases (Figure 1). In British Columbia, most affected was the Mission Medium Security Institution, a 1-hour drive away from Vancouver (64 cases, 30% prevalence) (Figure 1). Ontario’s Grand Valley Institution for Women, located in Kitchener (1-hour drive away from Toronto) also experienced a smaller outbreak of 8 cases (6% prevalence).

In the five institutions with one or more cases, we found that testing was largely reactive, with the onset of concentrated testing efforts occurring after COVID-19 outbreaks had already established. An exception to this observation was Quebec’s Federal Training Center, which was the only facility recording negative tests before the observations of positive tests (Figure 3). Further, though institutions all recorded a gradual increase in the number of tests performed over time, the high proportion of positive tests throughout the study period suggests that these prisons likely did not implement more intensive, wide-spread testing throughout the prison populations once a case was observed. An exception to this, however, was Ontario’s Grand Valley Women’s Institution, which recorded a significant jump in testing once the first cases were confirmed, with a majority of tests returning negative since April 7, 2020.

**Figure 3:**
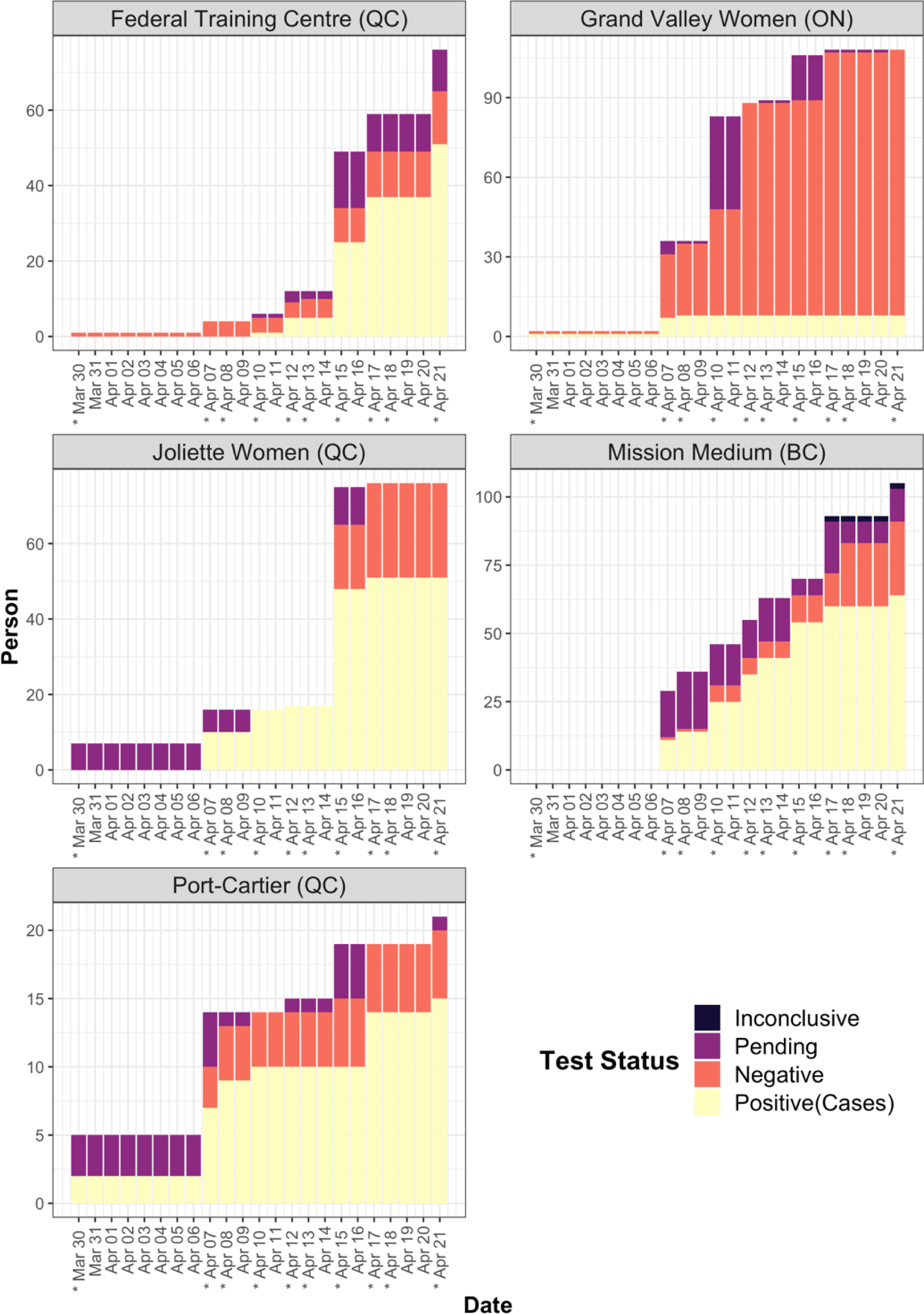
Testing patterns and outcomes between March 30 and April 21, 2020, in five penitentiaries with one or more recorded COVID-19 cases. * Indicated date at which data was reported by Correctional Services Canada.

### 3.3 Proportion of cases recovered and fatality inside versus outside prisons

The proportion of cases that had recovered inside federal penitentiaries ranged from 0% to 33% (Figure 1). The Susceptible, Infected, and Recovered plots (Figure 4) illustrate what proportion of prisoners each prison with one or more cases have become infected and have recovered or died throughout the study period. Of the five prisons with outbreaks, Quebec’s Joliette Women’s Institution and British Columbia’s Mission Medium Institution have the seen largest proportion of their population become infected and subsequently recover. Smaller proportions of cases have recovered in Quebec’s Federal Training Centre and Port-Cartier and Ontario’s Grand Valley Women’s Institution, and plot results suggest that a large proportion of prisoners remain susceptible in these institutions (Figure 4).

**Figure 4:**
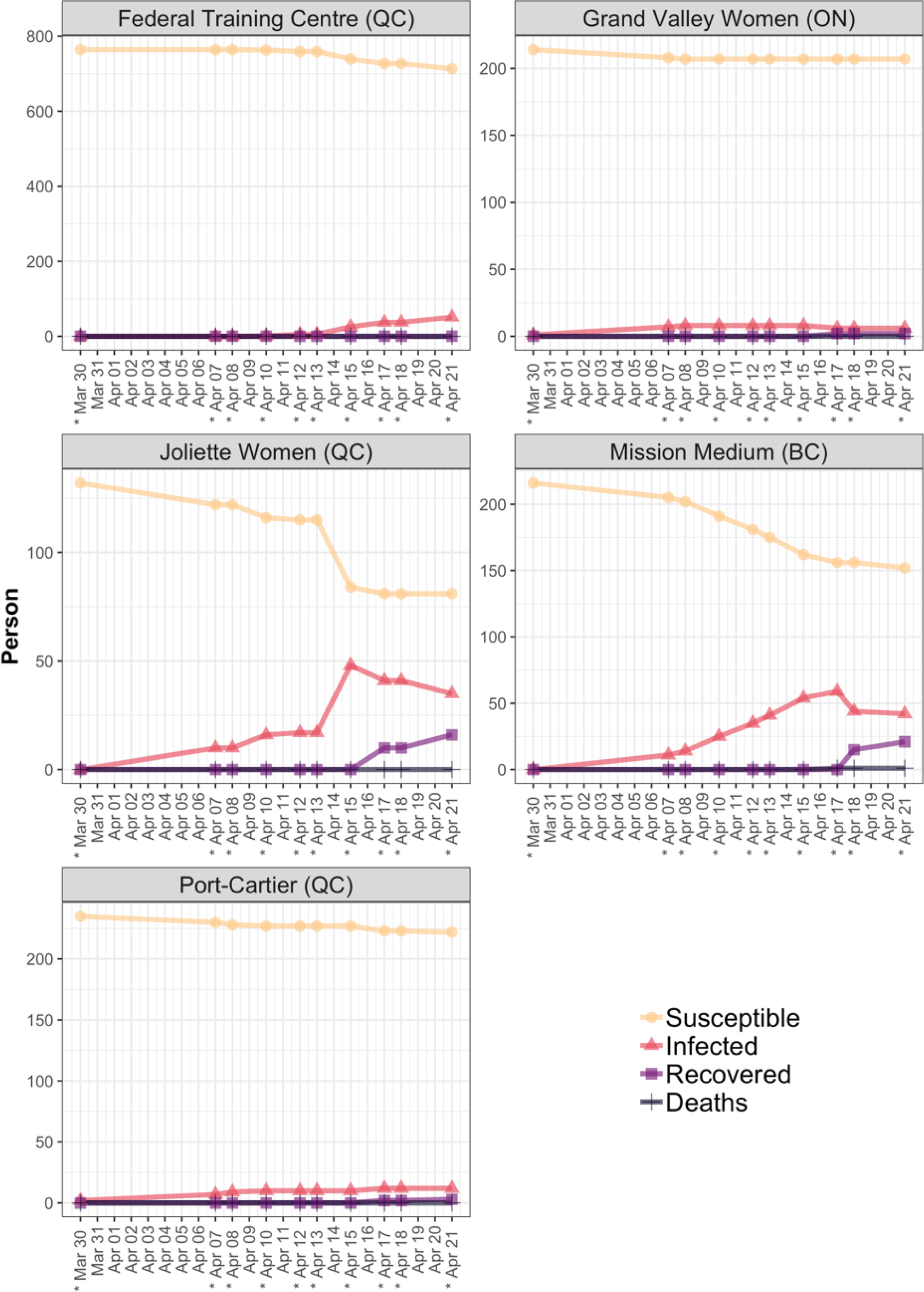
Number of susceptible, infected, recovered cases and deaths between March 30 and April 21, 2020, in penitentiaries with one or more recorded COVID-19 cases. * Indicates date at which data was reported by CSC.

In contrast to cases in the general population, a smaller proportion of cases in federal penitentiaries in Quebec, Ontario, and British Columbia had recovered (Figure 2). In both Ontario and British Columbia, the proportion of individuals who had recovered within prisons was half that of the proportion of cases recovered in the community. With the available evidence, it is difficult to know precisely what may be driving these differences in percent recovered inside versus outside of prisons. One explanation is that outbreaks inside prisons may have started more recently than those within the general public. Another possibility is that health and sanitary conditions within prisons may lead to lags in recovery. Further, this cross-sectional summary of differences must be interpreted with caution, since the proportion of cases that are recovered will vary according to the number of new incident cases observed in both settings.

At the time of data collection, one of the 189 laboratory-confirmed cases across all federal penitentiaries had died (0.5%), a 67% higher proportion than that observed in the general population, (0.3%, 1,966 of 614,340 cases). Case fatality estimates should be interpreted with caution, however, given the likely underestimation of the true number of cases (denominator value) in both settings.

### 3.4 Testing, prevalence and proportion recovered by sex

Among the 50 facilities included in these analyses, six are penitentiaries for women (Nova Institution for Women in Nova Scotia, Joliette Institution in Quebec, Grand Valley Institution for Women in Ontario, Edmonton Institution for Women in Alberta, Fraser Valley Institution in British Columbia, and Okimaw Ohci Healing Lodge in Saskatchewan). Together they hold up to 785 women (5% of the total capacity of 16,401 federal prisoners in Canada).

At the time of data collection, there were 59 cases of COVID-19 in women’s penitentiaries. These represented 31% of all cases in federal penitentiaries (N=189) (Figure 5), suggesting that women, and women’s penitentiaries, are over-represented among COVID-19 cases inside federal prisons. COVID-19 prevalence was 8 times higher among women’s prisons (8% prevalence) than prisons for men (1% prevalence) and 80 times higher than in the general Canadian population (0.1%).

**Figure 5:**
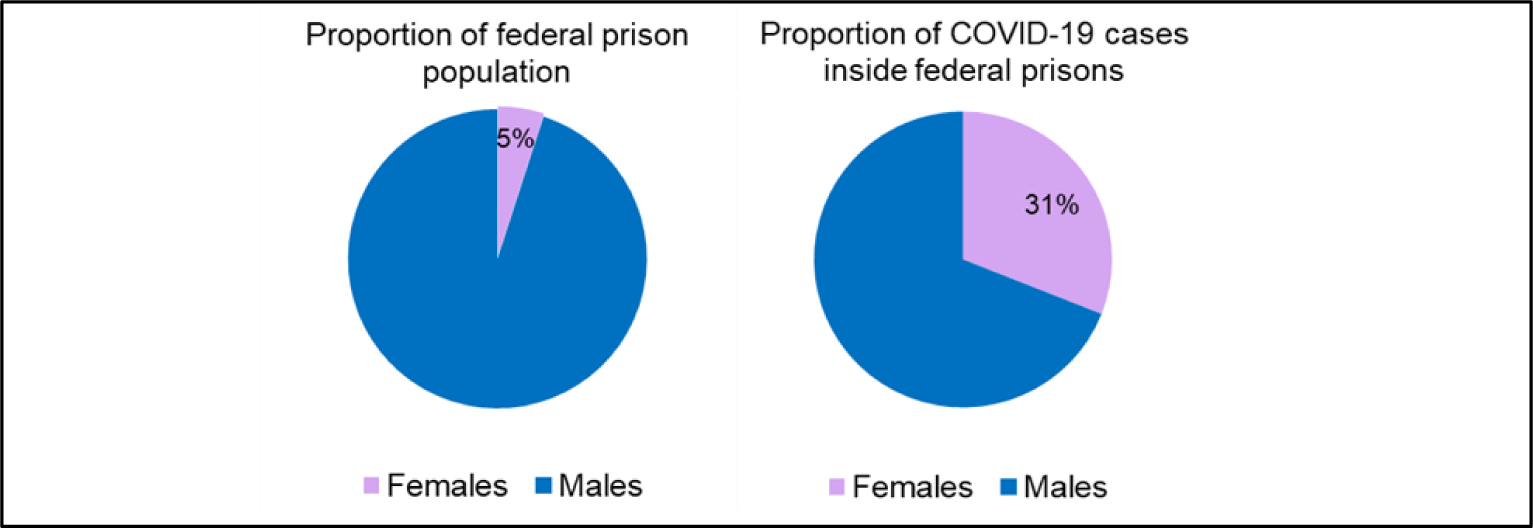
Distribution of women among federal prisons and prison COVID-19 cases (Canada, April 21, 2020).

## 4. DISCUSSION

### Key findings

The data suggest that at least five federal penitentiaries, one in British Columbia, three in Quebec and one in Ontario, are experiencing COVID-19 outbreaks. Each of these penitentiaries is reporting high levels of testing, compared to provincial and national rates, and this testing appears to be a reaction to the emergence of outbreaks that have already begun, rather than a proactive prevention effort. Though at least thirteen (26%) of facilities appear to be experiencing very low if any COVID-19 cases, these are also facilities that have reported low levels of testing per 1000 population, so it is unclear whether the low case counts reflect true case prevalence. We observed lower levels of testing than the Canadian general population average in 26 penitentiaries (72%) and a total absence of testing in 12 penitentiaries (24%). Our study also suggests a smaller proportion of cases inside prisons have recovered compared to cases in the general population. This may indicate that the spread of COVID-19 is lagging in penitentiaries, and the worst is yet to come. It may also indicate that prisoners face suboptimal conditions for recovery. We also found that at least three of the current penitentiary outbreaks are in proximity to city centers (e.g. Montreal and Vancouver), and higher COVID-19 prevalence has been observed in women’s (8% prevalence) compared to men’s prisons (1% prevalence), overall.

### Strengths and limitations

The primary strength of this study is its use of the best available data on COVID-19 testing and case incidence in Canadian Federal penitentiaries and the general Canadian population to give an early situational report on testing and infection-related outcomes within these facilities. This study is a baseline assessment from which future analyses using updated data can be based. A limitation of this study is our use of the maximum potential capacity of each penitentiary as the population denominator for rate calculations, rather than the ‘true’ population at the time of data collection. We expect that these denominators are likely too large, given that prisons may not be at their full capacity and pressures to release non-violent offenders to reduce the number of susceptible individuals within prisons [17]. This may have led to the under-estimation of testing and infection prevalence estimates. Further, this study was not able to also count the number of tests and cases identified within custodial and health staff within these same penitentiaries, which is an important contributing factor to disease transmission. While other deaths in federal penitentiaries were recorded by CSC during the study period (e.g. [18]), only one case was reported as COVID-19 related. It is unclear whether other deaths occurring within CSC facilities have or will be tested for COVID-19 post-mortem. Lastly, these findings may not be generalizable to provincial, remand, or immigration detention facilities, which may see far more movement in and out given the shorter duration of sentences.

### Implications

These early findings have several implications for public health and emergency outbreak response inside federal penitentiaries. The observation that at least five Canadian penitentiaries are experiencing outbreaks highlights the importance that all penitentiary populations and custodial and health care staff uphold rigorous prevention control practices (and be given the resources to do so), including regular hand hygiene and cleaning practices, the use of personal protection equipment, and physical distancing measures [19]. Scholars and legal experts have also emphasized the need to consider the release of prisoners to reduce the proportion of susceptible individuals within prisons [20].

Our findings of sparse testing across several penitentiaries in Canada suggests that a more proactive testing approach may be needed to help curb potential future outbreaks. This is especially the case given studies suggesting that up to 60% of COVID-19 cases may be asymptomatic [21–23]. Testing based on symptom-presence alone may thus need to be reconsidered within the confined spaces of prisons, as many cases may go undetected as long as testing is not expanded.

Relevant testing strategies could include universal testing or, active, sentinel-based testing. While universal testing [19] [24] is time and resource-intensive but is much more feasible within small confined populations than in the general public. In the context of COVID-19, this strategy has been used navy ships [21] and homeless shelters [25, 26] to identify all cases and adapt outbreak control interventions accordingly. In Canadian penitentiaries, universal testing may be most pertinent in prisons that have already reported confirmed cases. At the time of writing, British Columbia’s Mission Institution penitentiary — where one of the five outbreaks is unfolding — had reported planning universal testing of all inmates and staff [27]. However, none of the other institutions had indicated plans to use such an approach.

As an alternative to universal testing, active sentinel surveillance involves identifying a subset of prisons in which regular testing for COVID-19 among prisoners and staff, regardless of symptom presence, could be conducted. Test results would be used for monitoring in the months to come until a vaccine or effective treatment is identified. This approach may be most relevant in provinces with low numbers of new daily COVID-19 cases (e.g. currently New Brunswick, Nova Scotia, Saskatchewan and Manitoba currently) or in later in the pandemic, when community transmission subsides and testing all prisoners may be too costly and inefficient.

These types of proactive surveillance activities may be particularly important in the context of alternative options to reduce the spread of COVID-19, such as long-term cell-based confinement for the entire prison population. Confinement has been associated with severe mental health risk [28]—particularly for Indigenous and racialized populations [6]—and has come under considerable scrutiny in Canada in recent years [29].

Lastly, studies on the effectiveness of universal and sentinel-based testing strategies are warranted, as are mathematical models that explore the epidemiologic consequence of interventions (or lack thereof) within federal prisons on COVID-19 transmission, morbidity and death, as has been done in other country settings [30].

## 5. CONCLUSION

Gaps in COVID-19 testing in several jurisdictions and recorded outbreaks in several penitentiaries in Canada suggest that federal prisons represent an emerging battleground against the COVID-19 pandemic. With an ageing population that is particularly at high risk due to the prevalence of underlying chronic and infectious health conditions, prisoners represent a potentially neglected segment of society that is at high risk of bearing a high burden of COVID-19 related morbidity and mortality. As indigenous and racialized communities are over-represented within the Canadian prison system, COVID-19 prevention inside penitentiaries represents a pressing public health and health equity issue.

## Data Availability

Datasets analyzed and generated during this study are included in the supplementary materials.

## AUTHOR CONTRIBUTIONS

AB and AP conceptualized the study, with input from AS. AB and AP analyzed the data. AB and AP drafted the manuscript. AS responded to drafts of the manuscript.

## ETHICS COMMITTEE APPROVAL

None required

## 7. SUPPLEMENTARY FILE

### Federal Prison Population Data – as of April 21, 2020

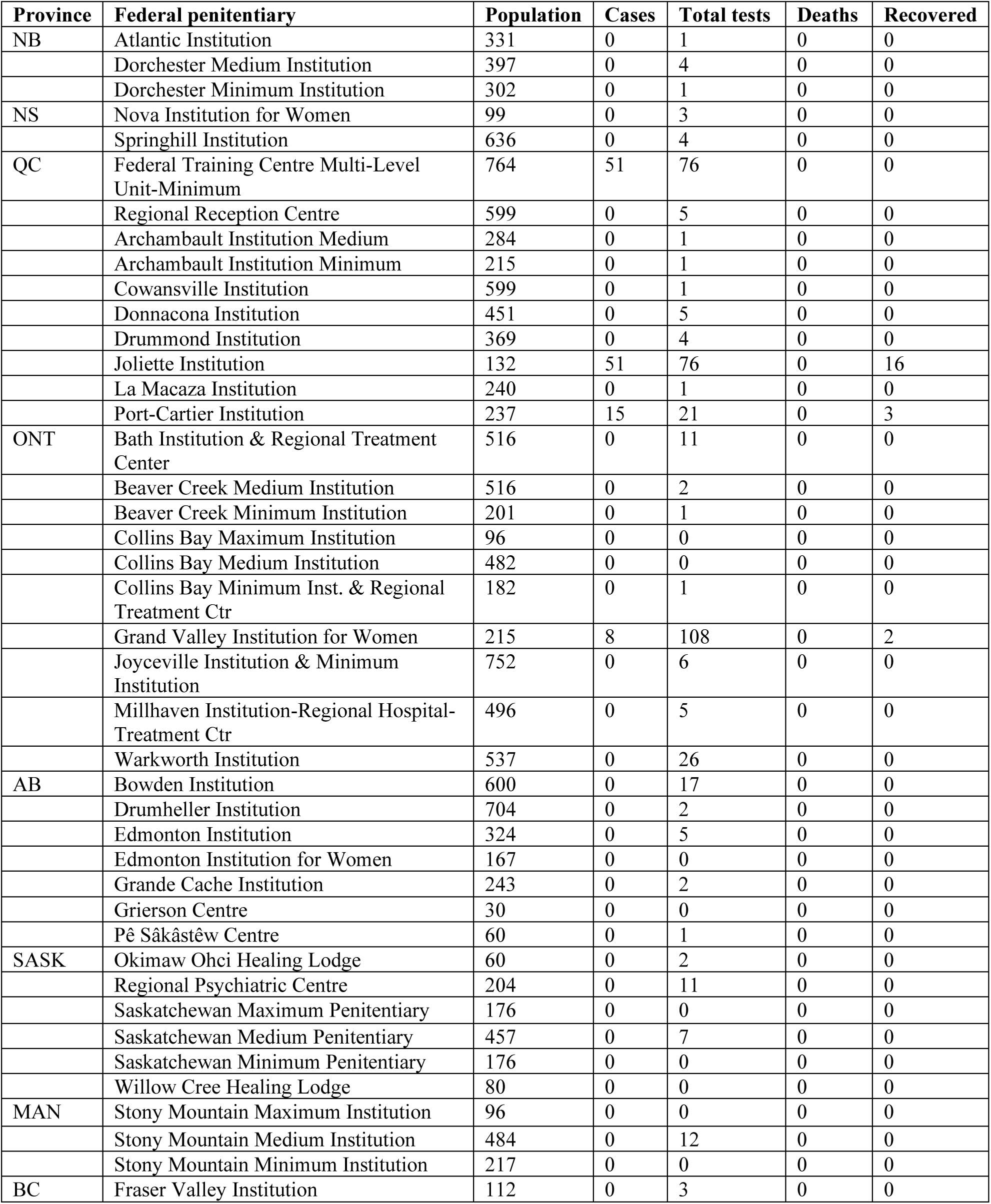
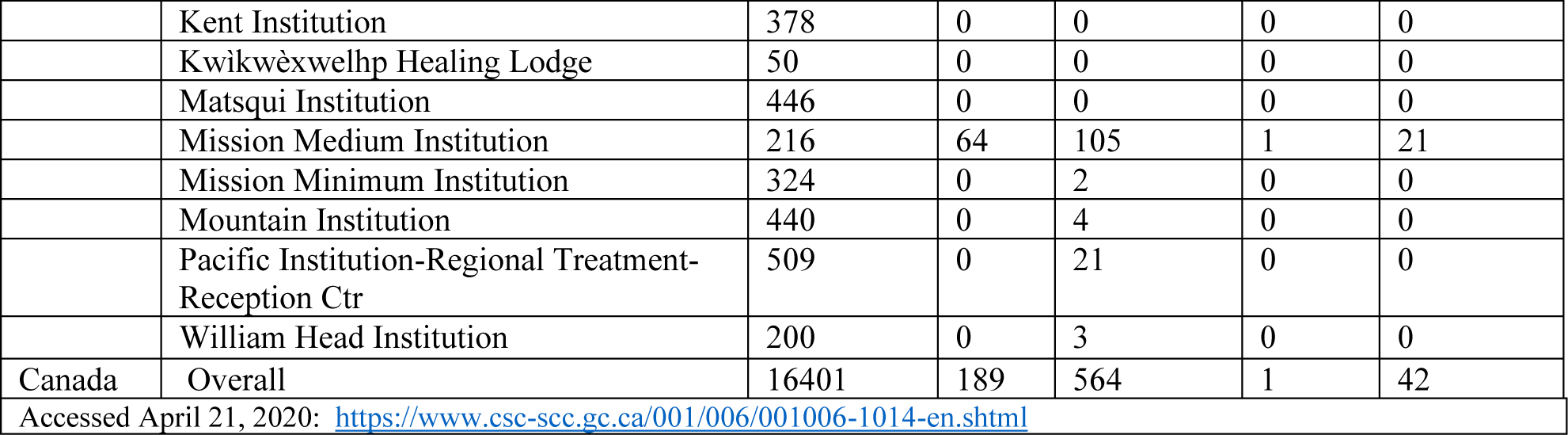

### General Population Data – as of April 21, 2020

| Province | Population | Positive tests | Total tests | Deaths | Recovered |
| --- | --- | --- | --- | --- | --- |
| New Brunswick | 779,993 | 118 | 11281 | 0 | 102 |
| Nova Scotia | 977,457 | 772 | 22927 | 12 | 330 |
| Quebec | 8,537,674 | 20965 | 174490 | 1134 | 4291 |
| Ontario | 14,711,827 | 12245 | 174170 | 659 | 6221 |
| Alberta | 4,413,146 | 3095 | 109015 | 61 | 1273 |
| Saskatchewan | 1,181,666 | 320 | 24811 | 4 | 252 |
| Manitoba | 1,377,517 | 255 | 20319 | 6 | 150 |
| British Columbia | 5,110,917 | 1724 | 65545 | 87 | 1041 |
| Canada (Overall) | 37,894,799 | 39805 | 614340 | 1966 | 13908 |
| Accessed April 21, 2020: http://openhub-esrica-apps.opendata.arcgis.com/datasets/covid-19-canada-1 ; Population data, accessed April 21, 2020: https://www150.statcan.gc.ca/t1/tb11/en/tv.action?pid=1710000901 |  |  |  |  |  |

### Data on facilities with one or more cases as of April 21, 2020

Data were available for March 30 and April 7-8, 10, 12, 13, 15, 17-18, 21 of 2020.

**Port Cartier Institution, Quebec** (Maximum Population Estimate: 237 prisoners)

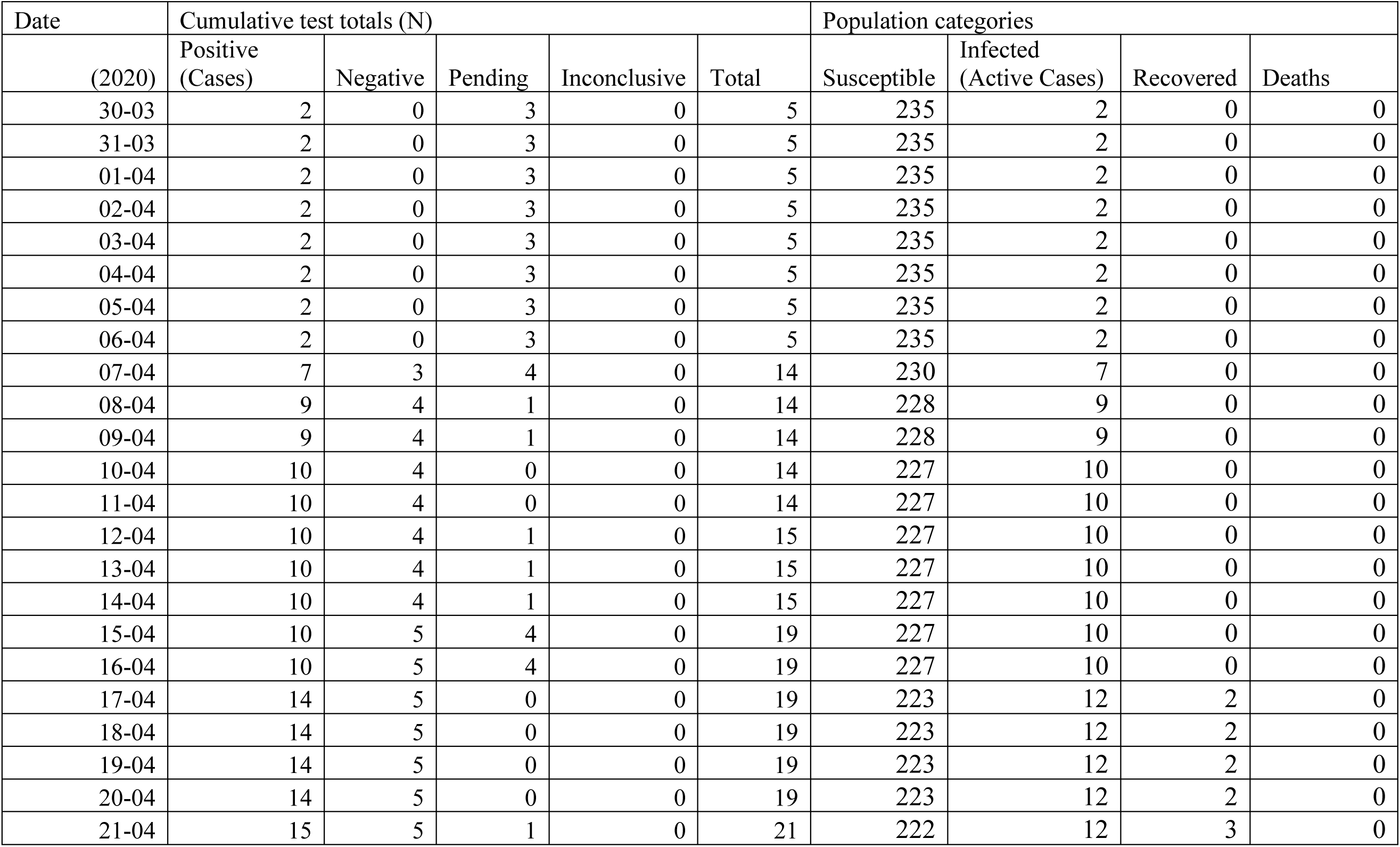

**Federal Training Center, Quebec** (Maximum Population Estimate: 764)

| Date | Cumulative test totals (N) |  |  |  |  | Population categories |  |  |  |
| --- | --- | --- | --- | --- | --- | --- | --- | --- | --- |
| (2020) | Positive (Cases) | Negative | Pending | Inconclusive | Total tests | Susceptible | Infected (Active Cases) | Recovered | Deaths |
| 30-03 | 0 | 1 | 0 | 0 | 1 | 764 | 0 | 0 | 0 |
| 31-03 | 0 | 1 | 0 | 0 | 1 | 764 | 0 | 0 | 0 |
| 01-04 | 0 | 1 | 0 | 0 | 1 | 764 | 0 | 0 | 0 |
| 02-04 | 0 | 1 | 0 | 0 | 1 | 764 | 0 | 0 | 0 |
| 03-04 | 0 | 1 | 0 | 0 | 1 | 764 | 0 | 0 | 0 |
| 04-04 | 0 | 1 | 0 | 0 | 1 | 764 | 0 | 0 | 0 |
| 05-04 | 0 | 1 | 0 | 0 | 1 | 764 | 0 | 0 | 0 |
| 06-04 | 0 | 1 | 0 | 0 | 1 | 764 | 0 | 0 | 0 |
| 07-04 | 0 | 4 | 0 | 0 | 4 | 764 | 0 | 0 | 0 |
| 08-04 | 0 | 4 | 0 | 0 | 4 | 764 | 0 | 0 | 0 |
| 09-04 | 0 | 4 | 0 | 0 | 4 | 764 | 0 | 0 | 0 |
| 10-04 | 1 | 4 | 1 | 0 | 6 | 763 | 1 | 0 | 0 |
| 11-04 | 1 | 4 | 1 | 0 | 6 | 763 | 1 | 0 | 0 |
| 12-04 | 5 | 4 | 3 | 0 | 12 | 759 | 5 | 0 | 0 |
| 13-04 | 5 | 5 | 2 | 0 | 12 | 759 | 5 | 0 | 0 |
| 14-04 | 5 | 5 | 2 | 0 | 12 | 759 | 5 | 0 | 0 |
| 15-04 | 25 | 9 | 15 | 0 | 49 | 739 | 25 | 0 | 0 |
| 16-04 | 25 | 9 | 15 | 0 | 49 | 739 | 25 | 0 | 0 |
| 17-04 | 37 | 12 | 10 | 0 | 59 | 727 | 37 | 0 | 0 |
| 18-04 | 37 | 12 | 10 | 0 | 59 | 727 | 37 | 0 | 0 |
| 19-04 | 37 | 12 | 10 | 0 | 59 | 727 | 37 | 0 | 0 |
| 20-04 | 37 | 12 | 10 | 0 | 59 | 727 | 37 | 0 | 0 |
| 21-04 | 51 | 14 | 11 | 0 | 76 | 713 | 51 | 0 | 0 |

**Joliette Women’s Institution, Quebec** (Maximum Population Estimate: 132 prisoners)

**Joliette Women's Institution, Quebec** (Maximum Population Estimate: 132 prisoners)
| Date | Cumulative test totals (N) |  |  |  |  | Population categories |  |  |  |
| --- | --- | --- | --- | --- | --- | --- | --- | --- | --- |
| (2020) | Positive (Cases) | Negative | Pending | Inconclusive | Total tests | Susceptible | Infected (Active Cases) | Recovered | Deaths |
| 30-03 | 0 | 0 | 7 | 0 | 7 | 132 | 0 | 0 | 0 |
| 31-03 | 0 | 0 | 7 | 0 | 7 | 132 | 0 | 0 | 0 |
| 01-04 | 0 | 0 | 7 | 0 | 7 | 132 | 0 | 0 | 0 |
| 02-04 | 0 | 0 | 7 | 0 | 7 | 132 | 0 | 0 | 0 |
| 03-04 | 0 | 0 | 7 | 0 | 7 | 132 | 0 | 0 | 0 |
| 04-04 | 0 | 0 | 7 | 0 | 7 | 132 | 0 | 0 | 0 |
| 05-04 | 0 | 0 | 7 | 0 | 7 | 132 | 0 | 0 | 0 |
| 06-04 | 0 | 0 | 7 | 0 | 7 | 132 | 0 | 0 | 0 |
| 07-04 | 10 | 0 | 6 | 0 | 16 | 122 | 10 | 0 | 0 |
| 08-04 | 10 | 0 | 6 | 0 | 16 | 122 | 10 | 0 | 0 |
| 09-04 | 10 | 0 | 6 | 0 | 16 | 122 | 10 | 0 | 0 |
| 10-04 | 16 | 0 | 0 | 0 | 16 | 116 | 16 | 0 | 0 |
| 11-04 | 16 | 0 | 0 | 0 | 16 | 116 | 16 | 0 | 0 |
| 12-04 | 17 | 0 | 0 | 0 | 17 | 115 | 17 | 0 | 0 |
| 13-04 | 17 | 0 | 0 | 0 | 17 | 115 | 17 | 0 | 0 |
| 14-04 | 17 | 0 | 0 | 0 | 17 | 115 | 17 | 0 | 0 |
| 15-04 | 48 | 17 | 10 | 0 | 75 | 84 | 48 | 0 | 0 |
| 16-04 | 48 | 17 | 10 | 0 | 75 | 84 | 48 | 0 | 0 |
| 17-04 | 51 | 25 | 0 | 0 | 76 | 81 | 41 | 10 | 0 |
| 18-04 | 51 | 25 | 0 | 0 | 76 | 81 | 41 | 10 | 0 |
| 19-04 | 51 | 25 | 0 | 0 | 76 | 81 | 41 | 10 | 0 |
| 20-04 | 51 | 25 | 0 | 0 | 76 | 81 | 41 | 10 | 0 |
| 21-04 | 51 | 25 | 0 | 0 | 76 | 81 | 35 | 16 | 0 |

**Mission Medium Security Institution** (Maximum Population Estimate: 216 prisoners)

| Date<br>(2020) | Cumulative test totals (N) |  |  |  |  | Population categories |  |  |  |
| --- | --- | --- | --- | --- | --- | --- | --- | --- | --- |
|  | Positive<br>(Cases) | Negative | Pending | Inconclusive | Total<br>tests | Susceptible | Infected (Active Cases) | Recovered | Deaths |
| 30-03 | 0 | 0 | 0 | 0 | 0 | 216 | 0 | 0 | 0 |
| 31-03 | 0 | 0 | 0 | 0 | 0 | 216 | 0 | 0 | 0 |
| 01-04 | 0 | 0 | 0 | 0 | 0 | 216 | 0 | 0 | 0 |
| 02-04 | 0 | 0 | 0 | 0 | 0 | 216 | 0 | 0 | 0 |
| 03-04 | 0 | 0 | 0 | 0 | 0 | 216 | 0 | 0 | 0 |
| 04-04 | 0 | 0 | 0 | 0 | 0 | 216 | 0 | 0 | 0 |
| 05-04 | 0 | 0 | 0 | 0 | 0 | 216 | 0 | 0 | 0 |
| 06-04 | 0 | 0 | 0 | 0 | 0 | 216 | 0 | 0 | 0 |
| 07-04 | 11 | 1 | 17 | 0 | 29 | 205 | 11 | 0 | 0 |
| 08-04 | 14 | 1 | 21 | 0 | 36 | 202 | 14 | 0 | 0 |
| 09-04 | 14 | 1 | 21 | 0 | 36 | 202 | 14 | 0 | 0 |
| 10-04 | 25 | 6 | 15 | 0 | 46 | 191 | 25 | 0 | 0 |
| 11-04 | 25 | 6 | 15 | 0 | 46 | 191 | 25 | 0 | 0 |
| 12-04 | 35 | 6 | 14 | 0 | 55 | 181 | 35 | 0 | 0 |
| 13-04 | 41 | 6 | 16 | 0 | 63 | 175 | 41 | 0 | 0 |
| 14-04 | 41 | 6 | 16 | 0 | 63 | 175 | 41 | 0 | 0 |
| 15-04 | 54 | 10 | 6 | 0 | 70 | 162 | 54 | 0 | 0 |
| 16-04 | 54 | 10 | 6 | 0 | 70 | 162 | 54 | 0 | 0 |
| 17-04 | 60 | 12 | 19 | 2 | 93 | 156 | 59 | 0 | 1 |
| 18-04 | 60 | 23 | 8 | 2 | 93 | 156 | 44 | 15 | 1 |
| 19-04 | 60 | 23 | 8 | 2 | 93 | 156 | 44 | 15 | 1 |
| 20-04 | 60 | 23 | 8 | 2 | 93 | 156 | 44 | 15 | 1 |
| 21-04 | 64 | 27 | 12 | 2 | 105 | 152 | 42 | 21 | 1 |

**Grand Valley Institution for Women** (Maximum Population Estimate: 112 prisoners)

| Date | Cumulative test totals (N) |  |  |  |  | Population categories |  |  |  |
| --- | --- | --- | --- | --- | --- | --- | --- | --- | --- |
| (2020) | Positive<br>(Cases) | Negative | Pending | Inconclusive | Total<br>tests | Susceptible | Infected<br>(Active Cases) | Recovered | Deaths |
| 30-03 | 1 | 1 | 0 | 0 | 2 | 214 | 1 | 0 | 0 |
| 31-03 | 1 | 1 | 0 | 0 | 2 | 214 | 1 | 0 | 0 |
| 01-04 | 1 | 1 | 0 | 0 | 2 | 214 | 1 | 0 | 0 |
| 02-04 | 1 | 1 | 0 | 0 | 2 | 214 | 1 | 0 | 0 |
| 03-04 | 1 | 1 | 0 | 0 | 2 | 214 | 1 | 0 | 0 |
| 04-04 | 1 | 1 | 0 | 0 | 2 | 214 | 1 | 0 | 0 |
| 05-04 | 1 | 1 | 0 | 0 | 2 | 214 | 1 | 0 | 0 |
| 06-04 | 1 | 1 | 0 | 0 | 2 | 214 | 1 | 0 | 0 |
| 07-04 | 7 | 24 | 5 | 0 | 36 | 208 | 7 | 0 | 0 |
| 08-04 | 8 | 27 | 1 | 0 | 36 | 207 | 8 | 0 | 0 |
| 09-04 | 8 | 27 | 1 | 0 | 36 | 207 | 8 | 0 | 0 |
| 10-04 | 8 | 40 | 35 | 0 | 83 | 207 | 8 | 0 | 0 |
| 11-04 | 8 | 40 | 35 | 0 | 83 | 207 | 8 | 0 | 0 |
| 12-04 | 8 | 80 | 0 | 0 | 88 | 207 | 8 | 0 | 0 |
| 13-04 | 8 | 80 | 1 | 0 | 89 | 207 | 8 | 0 | 0 |
| 14-04 | 8 | 80 | 1 | 0 | 89 | 207 | 8 | 0 | 0 |
| 15-04 | 8 | 81 | 17 | 0 | 106 | 207 | 8 | 0 | 0 |
| 16-04 | 8 | 81 | 17 | 0 | 106 | 207 | 8 | 0 | 0 |
| 17-04 | 8 | 99 | 1 | 0 | 108 | 207 | 6 | 2 | 0 |
| 18-04 | 8 | 99 | 1 | 0 | 108 | 207 | 6 | 2 | 0 |
| 19-04 | 8 | 99 | 1 | 0 | 108 | 207 | 6 | 2 | 0 |
| 20-04 | 8 | 99 | 1 | 0 | 108 | 207 | 6 | 2 | 0 |
| 21-04 | 8 | 100 | 0 | 0 | 108 | 207 | 6 | 2 | 0 |

